# Magnitude, change over time, demographic characteristics and geographic distribution of excess deaths among nursing home residents during the first wave of COVID-19 in France: a nationwide cohort study

**DOI:** 10.1101/2021.01.09.20248472

**Authors:** Florence Canouï-Poitrine, Antoine Rachas, Martine Thomas, Laure Carcaillon-Bentata, Roméo Fontaine, Gaëtan Gavazzi, Marie Laurent, Jean-Marie Robine, on behalf of the COMONH consortium

**Affiliations:** Univ Paris Est Creteil, Inserm, IMRB U955, CEpiA Team, F-94000, Creteil, France; APHP, Henri-Mondor Hospital, Public Health Department, F-94000, Creteil, France; Cnam, Direction de la Stratégie, des Etudes et des Statistiques, F-75000, Paris, France; Santé Publique France (SpF), F-94410, Saint-Maurice, France; INED, Mortality, Health and Epidemiology (UR5), F-93300, Aubervilliers, France; Grenoble Alpes University Hospital, Geriatric Department, F-38000, Grenoble, France; University of Grenoble-Alpes, GREPI TIMC-IMAG, CNRS UMR 552, F-38000, Grenoble, France; APHP, Henri-Mondor Hospital, Geriatric Department, F-94000 Creteil, France; Univ Paris, INSERM, CNRS, EHSS, CERMES3, F-75000, Paris, France; Univ Montpellier, EPHE, INSERM, MMDN, F-34000, Montpellier, France; PSL Research University, F-75000, Paris, France

**Keywords:** COVID-19, Excess deaths, Elderly adults, Nursing home

## Abstract

**Importance:** Nursing home (NH) residents are particularly vulnerable to SARS-CoV-2 infections and coronavirus disease 2019 (COVID-19) lethality. However, excess deaths in this population have rarely been documented.

**Objectives:** The primary objective was to assess the number of excess deaths among NH residents during the first wave of the COVID-19 pandemic in France. The secondary objectives were to determine the number of excess deaths as a proportion of the total excess deaths in the general population and determine whether a harvesting effect was present.

**Design:** We studied a cohort of 494,753 adults (as of March 1^st^, 2020) aged 60 and over in 6,515 NHs in mainland France. This cohort was exposed to the first wave of the COVID-19 pandemic (from March 1^st^ to May 31^st^, 2020) and was compared with the corresponding, reference cohorts from 2014 to 2019 (using data from the French National Health Data System).

**Main outcome and measures:** The main outcome was all-cause death. Weekly excess deaths and standardized mortality ratios (SMRs) were estimated.

**Result:** There were 13,505 excess deaths among NH residents. Mortality increased by 43% (SMR: 1.43). The mortality excess was higher among males than among females (SMR: 1.51 and 1.38, respectively) and decreased with age (SMRs in females: 1.61 in the 60-74 age group, 1.58 for 75-84, 1.41 for 85-94, and 1.31 for 95 or over; Males: SMRs: 1.59 for 60-74, 1.69 for 75-84, 1.47 for 85-94, and 1.41 for 95 or over). We did not observe a harvesting effect (up until August 30^th^, 2020). By extrapolating to all NH residents nationally (N=570,003), the latter accounted for 51% of the total excess deaths in the general population (N=15,114 out of 29,563).

**Conclusion:** NH residents accounted for about half of the total excess deaths in France during the first wave of the COVID-19 pandemic. The excess death rate was higher among males than females and among younger residents than among older residents. We did not observe a harvesting effect. A real-time mortality surveillance system and the identification of individual and environmental risk factors might help to design the future model of care for older dependent adults.

**Key points:**

- During the first wave of the COVID-19 pandemic in France, the mortality among nursing home residents increased by 43%.
- Nursing home residents accounted for 51% of the total excess deaths in France.
- The excess mortality was higher among younger residents than among older residents.
- The excess mortality was higher among males than among females.
- We did not observe a harvesting effect during the study period (ending on August 30^th^, 2020, i.e., three months after the end of the first wave).

## Introduction

In high-income countries, almost all deaths cause by coronavirus disease 2019 (COVID-19) have occurred in older adults. In the United States, 80% of the deaths occurred among people aged 65 or over. ^1,2^ In France, 71% of the in-hospital deaths between March 1^st^ and June 2^nd^, 2020, concerned patients aged 75 or over.^1^ In China, the case fatality rate was 2% overall, 8% in patients aged 70 to 79, and 15% in patients aged 80 and over.^3^ In a meta-analysis of 58 studies covering 122,191 hospitalized patients worldwide, COVID-19 lethality was more strongly associated with age over 65 than with comorbidities, obesity, hypertension, diabetes, cardiovascular disease, or cancer.^4,5^

Soon after the start of the pandemic, there were concerns about nursing homes (NHs) as hotspots of contagion and deaths.^6^ Several European governments were criticized for reporting only in-hospital deaths because it soon became clear that deaths in NHs accounted for a nonnegligible proportion of deaths-between 25% and 66% in the United States and in European countries. ^7-9^ Accordingly, the surveillance systems built to count daily COVID-19 deaths started to distinguish between hospitals, NHs, and private homes. ^10,11^ However, these systems give a fragmented view of the overall impact of the pandemic on NH residents, since some of the latter died in their NH and others died in hospital.

Hence, reporting COVID-19-related deaths alone gives a partial view of the pandemic’s impact. In contrast, the estimation of excess deaths takes account of the pandemic’s negative (mortality-increasing) and positive (mortality-decreasing) direct and indirect effects.^12,13^

We hypothesized that the overall impact of the first wave of the COVID-19 pandemic on NH residents in France could be assessed by leveraging the national NH administrative database (RESID-EHPAD, part of the French National Health Data System).^14^ The primary objective of the present study was to assess the magnitude of excess mortality among NH residents in France during the first wave of the COVID-19 pandemic overall and by age, sex and territorial unit. The secondary objectives were to (i) determine excess deaths in NHs as a proportion of total excess deaths, (ii) assess changes over time of excess mortality, and determine whether a harvesting effect (i.e., a short-term forward shift in mortality^15,16^) was present.

## Methods

### Design

This was a nationwide cohort study of NH residents exposed (the 2020 cohort) or not (the 2014-2019 cohorts) to the first wave of the COVID-19 pandemic in France. The six-year reference period was used to smooth out annual variations in mortality.^12,15^

### Setting

Residential homes for dependent older adults requiring nursing and medical care (*“établissements d’hébergement pour personnes âgées dépendantes”*) located throughout mainland France (the COVID-19 pandemic occurred later in France’s overseas territories).

### Participants

The study cohorts were composed of adults aged 60 or over residing in an NH on March 1^st^ of each year. Residents admitted to an NH between March 1^st^ and December 31^st^ were not included.

### Follow-up

Two analysis periods were defined. The first period (from March 1^st^ to May 31^st^, 2020) covered the first wave of the COVID-19 pandemic in France and was compared with the same period in the previous 6 years (2014-2019). The second period ran from December 30^th^, 2019, to August 30^th^, 2020. Each week in this period was compared with the same week in the previous 6 years.^17^

### Data sources

RESID-EHPAD is a French national online database in which all admissions and discharges of NH residents are reported. NHs funded and managed by the French national health insurance fund for agricultural workers (*Mutualité Sociale Agricole*) do not contribute to RESID-EHPAD. Although the RESID-EHPAD database is part of the French National Health Data System (*Système National des Données de Santé*, SNDS), it cannot be accessed routinely by researchers. ^14,18^ The SNDS contains extensive individual data on all expenditure covered by France’s mandatory public health insurance system, together with the dates of deaths provided by the French National Statistics Office (*Institut National de la Statistique et des Etudes Economiques*, INSEE) through local health insurance offices. The standard, anonymized SNDS data extracts made available to authorized researchers do not contain the RESID-EHPAD variables. Analyses of French National Health Data System were performed with the permission of the independent French data protection authority (Commission Nationale Informatique et Libertes) and by decree (articles R. 1461‐12 and seq. of the French Public Health Code).

### Outcome

The main outcome was death, regardless of where it reportedly occurred.

### Variables

We analyzed sex, age, and the NH’s geographic location. Four age groups were pre-specified: 60-74, 75-84, 85-94, and 95 or over. Geographic locations were classified according to the third level of the Nomenclature of Territorial Units for Statistics (NUTS 3), corresponding to the *département* (of which there are 96 in mainland France).^19^

### Statistical analysis

The baseline characteristics of the NHs and their residents were described as the frequency (percentage) or the mean (standard deviation (SD)). Two indicators were used to summarize observed mortality, the total number of deaths, and the cumulative mortality rates over the whole period and on a weekly basis. For calculation of the mortality rates, the observation time per person present at the beginning of period (i.e., present on March 1^st^ or at the beginning of a specific week) was calculated up to the date of death if this event occurred till the end of the period.

Firstly, expected numbers of deaths were estimated as counterfactual values by applying the 5-year age group- and sex-specific mortality rates observed for the 2014-2019 cohorts to the 2020 cohort. Secondly, numbers of excess deaths were calculated as observed deaths minus expected deaths for the first-wave period and per week. Thirdly, standardized mortality ratios (SMRs) were obtained by computing the ratio between observed and expected deaths.^17^ Excess mortality patterns were studied according to age, sex and geographical (NUTS3) location. Lastly, to estimate the excess death among the whole NH population, the SMR was applied to missing NH residents from the same NUTS3 unit. Data on the numbers of missing NH residents were obtained from the French national registry of healthcare and social establishments (*Fichier national des établissements sanitaires et sociaux*, FINESS) as of March 29^th^, 2020. We then computed excess deaths among NH residents as a proportion of those in the whole population (according to data from INSEE) for 2020 vs. 2014-2019. ^20^ Statistical analyses were performed with SAS Enterprise Guide software (version 4.3, SAS Institute Inc. Cary, NC). Results were reported according to the Strengthening the Reporting of Observational Studies in Epidemiology guidelines.

## Results

### Baseline characteristics

The 2020 cohort included 494,753 residents from 6515 NHs in mainland France. The baseline characteristics of the residents and NHs are summarized in Table 1. 74% of the residents were female. The mean (SD) age was 89 (8) years for females and 84 (9) years for males. The mean number of residents per NH was 83 (43).

**Table 1:** Baseline characteristics of the residents and the NHs.

| Table 1 : Baseline characteristics of the residents and the NHs |  |  |  |
| --- | --- | --- | --- |
| Variable | Item | n (%) or mean (SD) |  |
| **Residents** |  | **N=494,753** |  |
|  |  | Females | Males |
| Sex |  | 368,165 (74) | 126,588 (26) |
| Age (year), mean (SD) |  | 89 (8) | 84 (9) |
| Age groups (years) | 60-74 | 22,755 (6) | 24,099 (19) |
|  | 75-84 | 60,376 (16) | 30,957 (25) |
|  | 85-94 | 201,898 (55) | 57,161 (45) |
|  | 95 or over | 83,136 (23) | 14,371 (11) |
| **NHs** |  | **N=6515** |  |
| For-profit |  | 3797 (58) |  |
| Number of residents, mean (SD) |  | 83 (43) |  |
| Number of residents (classes) | <50 | 892 (14) |  |
|  | 50-74 | 1898 (29) |  |
|  | 75-99 | 2442 (37) |  |
|  | 100-149 | 917 (14) |  |
|  | 150 or more | 366 (6) |  |
| NH: nursing home |  |  |  |

### Mortality rates and excess deaths

A total of 44,666 NH residents died between March 1^st^ and May 31^st^, 2020, versus an average of 30,647 in the reference cohorts. The cumulative mortality rate in NHs during the first wave was 9.5%, versus 6.4% during the same period in the previous 6 years. In all age groups, the mortality rate was higher in males than in females (Table 2). The mortality rate increased with age (Table 2). There were 13,505 excess deaths among NH residents, giving an SMR of 1.43; hence, mortality increased by 43% in 2020, relative to the previous 6 years. Overall, there were 570,003 NH residents in France on March, 2020. Applying the SMR for NH residents in the same NUTS3 to the missing residents, we estimated that there were 15,114 excess deaths among the entire population of NH residents. There were 29,563 excess deaths in the general population in 2020, relative to 2014-2019. Therefore, NH residents may have accounted for 51% of the excess deaths in the France’s population.

**Table 2:** Mortality during the first wave^a^ of the COVID-19 epidemic in France, by sex and by age group.

|  |  |  |  |  |  |  |
| --- | --- | --- | --- | --- | --- | --- |
| sex | age group (years) | number of residents | number of deaths | cumulative mortality rate (%)<sup>b</sup> | number of excess deaths | SMR<sup>c</sup> |
| both | all | 494 753 | 44 666 | 9.5 | 13,505 | 1.43 |
| males | all | 126 588 | 14 445 | 12.1 | 4905 | 1.51 |
|  | 60-74 | 24 099 | 1 180 | 5.0 | 437 | 1.59 |
|  | 75-84 | 30 957 | 3 132 | 10.7 | 1282 | 1.69 |
|  | 85-94 | 57 161 | 7 680 | 14.5 | 2473 | 1.47 |
|  | 95 or over | 14 371 | 2 453 | 18.8 | 714 | 1.41 |
| females | all | 368 165 | 30 221 | 8.6 | 8599 | 1.38 |
|  | 60-74 | 22 755 | 851 | 3.8 | 322 | 1.61 |
|  | 75-84 | 60 376 | 3 606 | 6.2 | 1325 | 1.58 |
|  | 85-94 | 201 898 | 16 230 | 8.4 | 4684 | 1.41 |
|  | 95 or over | 83 136 | 9 534 | 12.2 | 2268 | 1.31 |
| <sup>a</sup> between March 1st and May 31st, 2020 |  |  |  |  |  |  |
| <sup>b</sup> number of deaths divided by the number of person-times between March 1st and May 31st, 2020. |  |  |  |  |  |  |
| <sup>c</sup> SMR: standardized mortality ratio |  |  |  |  |  |  |

### Changes over time in the death rate

The observed number of deaths per week exceeded the expected number of deaths from week 11 to week 26, with a peak in weeks 14 (5,486 deaths) and 15 (5,474 deaths) (Figure 1.A). The weekly mortality rate reached 1.16% during week 15. More than 3,000 excess deaths per week were observed in weeks 14 and 15 (SMR: 2.37) (Figure 1 C,D). We did not observe a lack of deaths (i.e., a harvesting effect) at any time during the study period (ending in week 35, August 2020; Figure 1).

**Figure 1:**
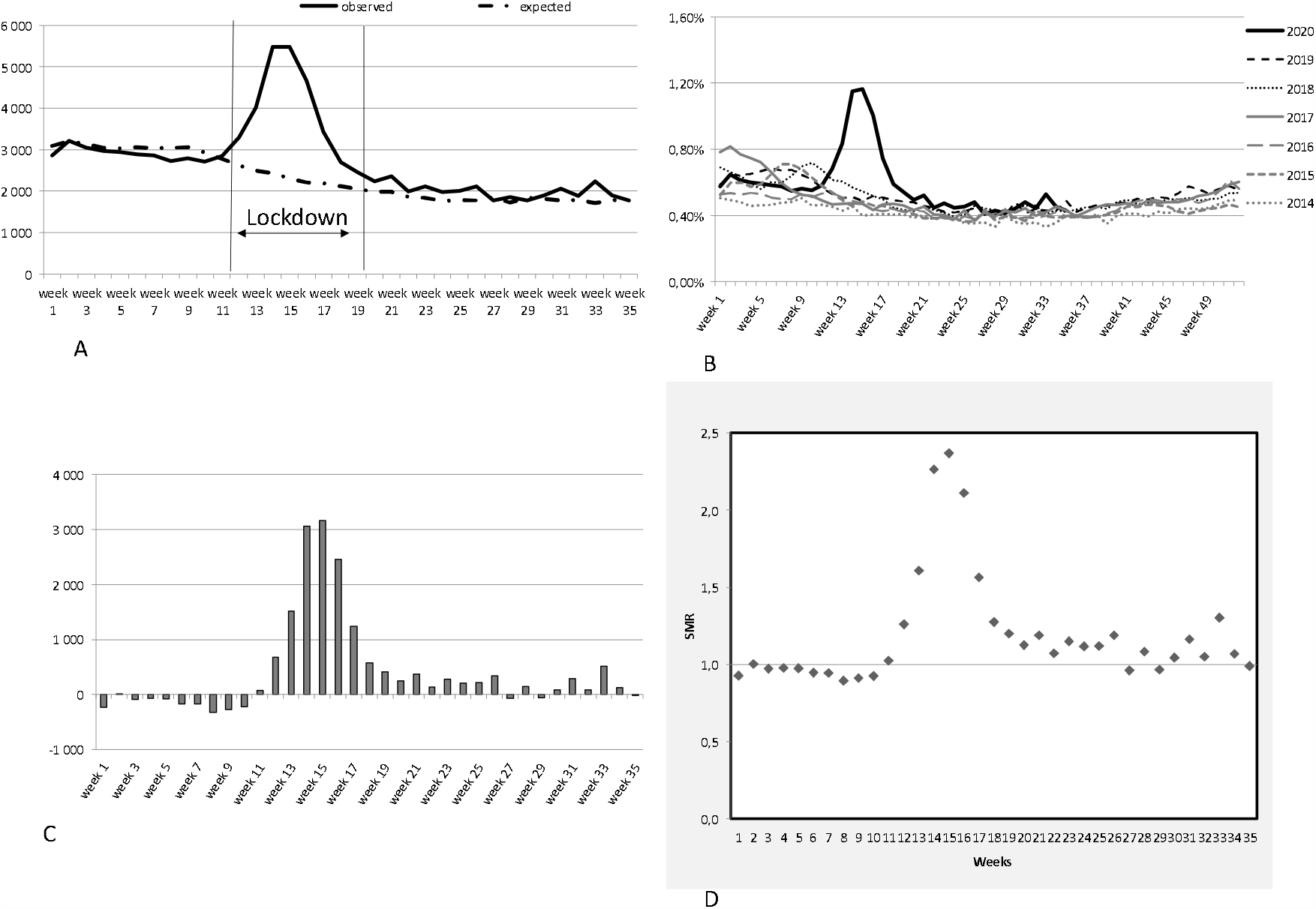
the weekly observed and expected deaths (A), mortality rates (B), weekly excess deaths (C), and standardized mortality ratios (D)

### Age- and sex-specific mortality rates and excess deaths

The changes over time were similar in the four age groups and in both sexes (Figure 3). The excess mortality rate was higher among males than among females (Table 2) and decreased with age in females (Table 2 and Figure 2). In males, the SMR was higher in the 75-84 age group than in the 60-74 age group and then decreased again for older age groups (Table 2 and Figure 3).

**Figure 2:**
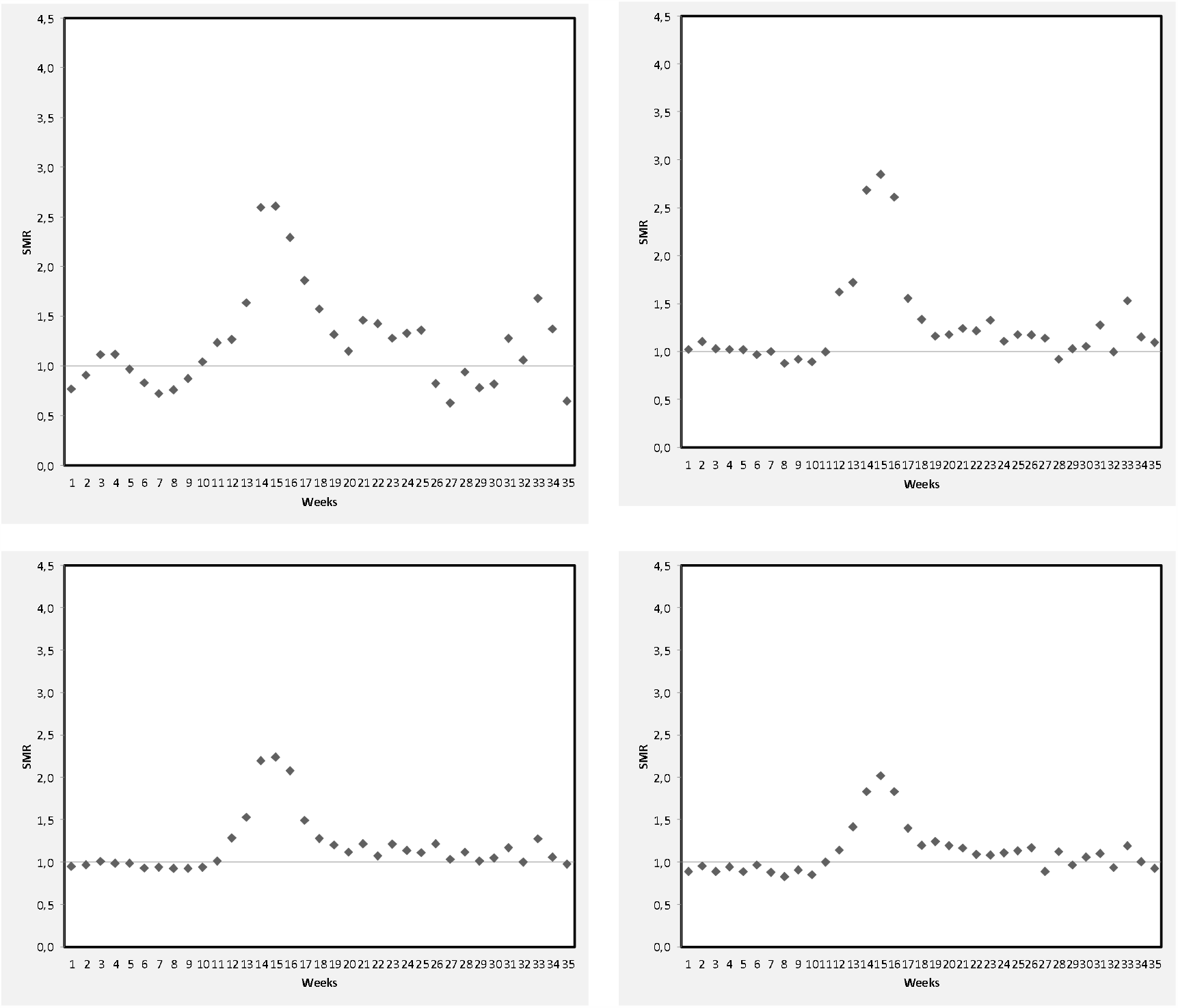
the weekly standardized mortality ratio in females aged 60-74 (A), 75-84 (B), 85-94 (C), and 95 or over (D)

**Figure 3:**
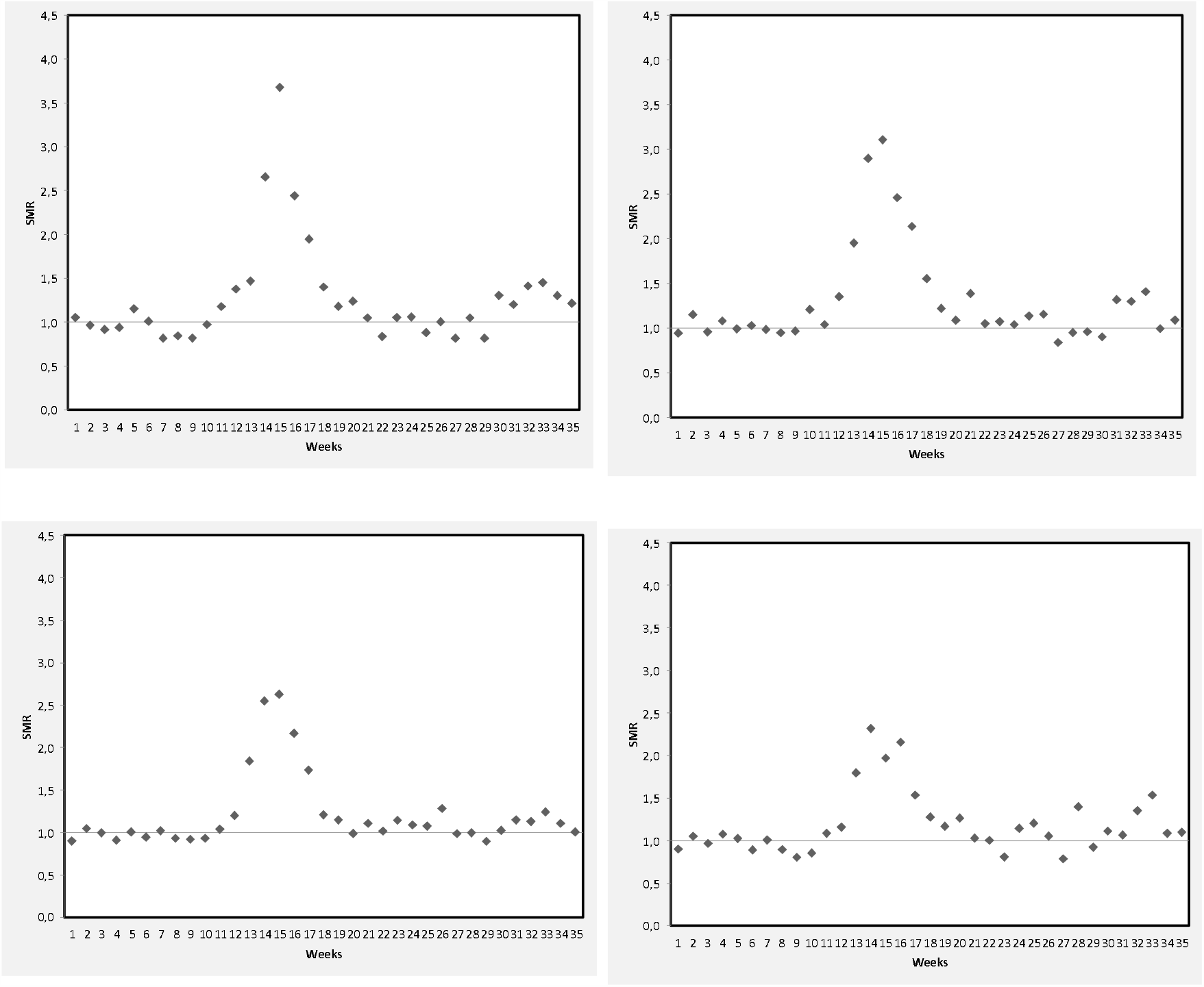
the weekly standardized mortality ratio in males aged 60-74 (A), 75-84 (B), 85-94 (C), and 95 or over (D)

### Geographic distribution of mortality rates and excess deaths

At the NUTS3 level, the mortality rate among NH residents during the first wave of the pandemic ranged from 5.3% in Lozère (a rural *département* with the lowest population density) to 22.2% in the socially deprived Paris suburb of Seine-Saint-Denis. Accordingly, the SMR ranged from 0.92 in Lozère to 3.47 in Seine-Saint-Denis (Figure 4).

**Figure 4:**
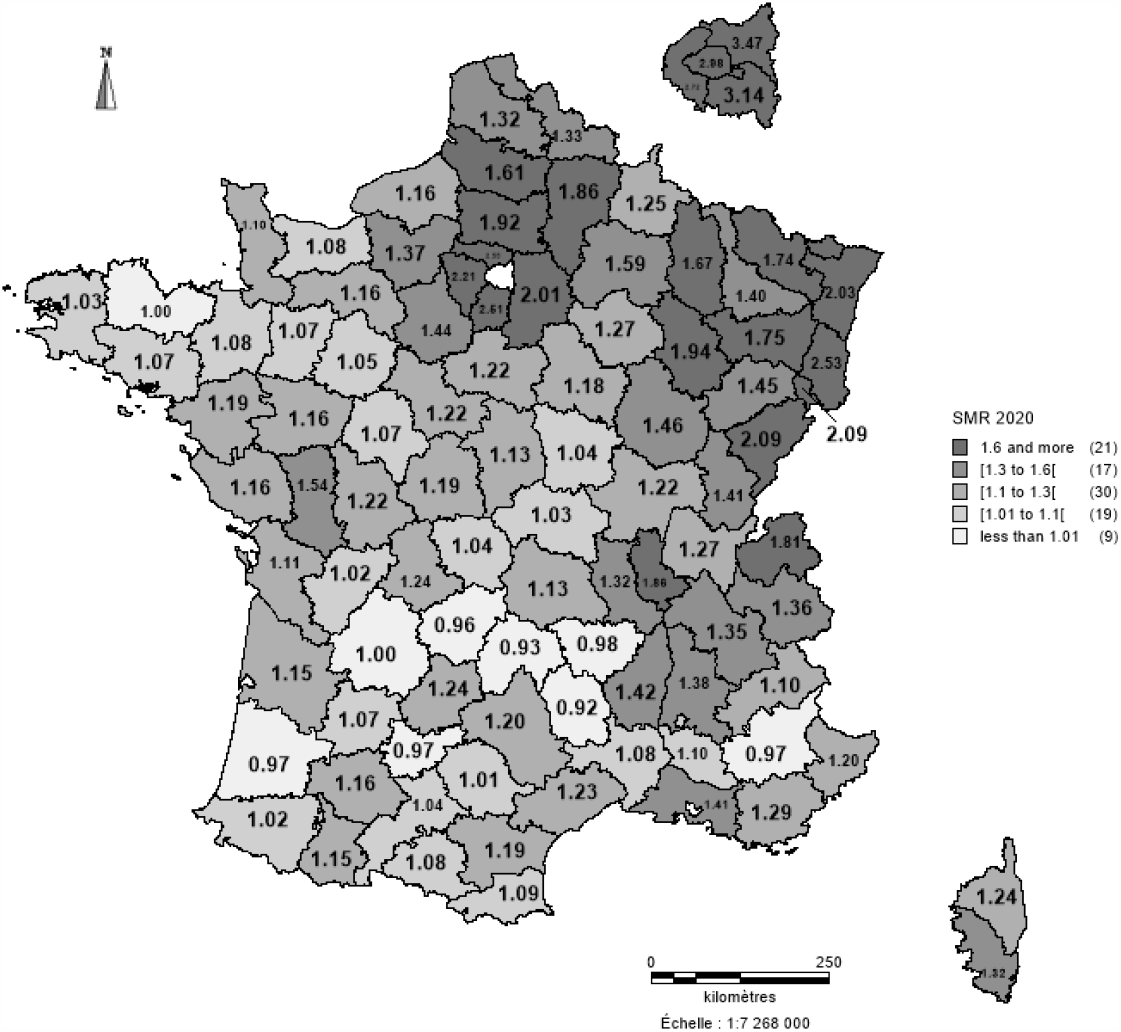
Standardized mortality ratios per level 3 territorial unit (defined according to the Nomenclature of Territorial Units for Statistics).

## Discussion

Among 494,753 residents of 6,515 NHs in mainland France, we observed 13,505 excess deaths during the first wave of the COVID-19 pandemic (March 1^st^ to May 31^st^, 2020), relative to the previous 6 years. The mortality increased by 43%. The excess mortality was higher in males than in females (51% and 38%, respectively) and decreased with age. The changes over time in excess mortality were similar in all age groups and in both sexes. At the peak of the first wave (weeks 14 and 15), there were more than 3,000 excess deaths (giving SMRs of 2.26 and 2.37, respectively). The excess mortality varied greatly from one territorial unit to another (from −8% to +247%). We did not observe a harvesting effect at any time during the study period, which ending in week 35, August 30^th^, 2020 (i.e., 3 months after the end of the first wave). The NH residents accounted for 51% of the excess mortality in the general population during the first wave of the COVID-19 pandemic.

To the best of our knowledge, the present study is the first to have assessed the overall impact of the COVID-19 pandemic on deaths of NH residents. As mentioned above, reporting at the start of the health crisis typically included in-hospital deaths but not deaths in NHs, and only the place of death (rather than the place of residence at the time of death) was taken into account.^10,11^ In England & Wales, the United Kingdom’s Office for National Statistics reported 57,074 excess deaths between March and May 2020, of which 26,211 (46%) occurred in care homes.^10,11^ In France, the INSEE estimated that the number of deaths in NHs increased by 54% between March and April 2020, compared with the previous year. However, these figures did not take account of NH residents who died after being transferred to a hospital. Moreover, there was a degree of uncertainty regarding the reporting on the place of death by municipal offices of vital records.^10^

The majority of studies in the literature reported on COVID-19-related deaths rather than excess deaths. In France and in England & Wales, respectively 10,264 and 9,039 COVID-19-related deaths in NHs were reported between March and May 2020. Furthermore, 3,615 and 3,444 in-hospital COVID-19-related deaths of NH residents were respectively reported. ^1,11^ A lack of PCR tests at the beginning of the pandemic, atypical clinical presentations in older people, reports of both suspected and confirmed cases to the health surveillance system, and (in France) erroneous reporting of the place of death by municipal offices of vital records all led to uncertainty regarding deaths attributed to COVID-19 in NHs.^21,22^

The steep increase and then decrease in excess deaths during the first wave were similar to those observed in 65 NHs in New York City.^23^ In France, however, the decrease in excess deaths began 4 weeks after visits with NH residents were prohibited (week 11) and became much more pronounced 3 weeks after the start of the nationwide period of lockdown (week 12). The mortality rate among NH residents in New York City decreased 5 weeks after visits were prohibited.^23^

We also found an inverse relationship between the relative magnitude of excess deaths and increasing age in females. In males, the relative excess death rate was consistently lower in older age groups than in younger age groups but the relationship was not linear: the relative number of excess deaths was higher in the 75-84 age group than in the 60-74 age group. Similar results have been observed in the general population. ^22^ In Italy, for instance, Conti and colleagues found a lower excess mortality in nonagenarians than in their younger counterparts.^24^ This might have been due to a healthy survivor effect. Younger NH residents might have more comorbidities-notably stroke and diabetes, which are risk factors for COVID-19 severity and lethality -than older residents. ^5,24-26^ Interestingly, a multicenter retrospective cohort study of 821 older COVID-19 patients (average age of 86 years) hospitalized in acute geriatric ward did not evidence an age-group-related difference in mortality after adjustment for comorbidities and dependency.^27^ The excess mortality was higher among males than among females; this is in line with the well-known excess mortality among males in other settings ^25^. This sex difference might also be due to the clinical profile of males in NHs, who are younger but have more cardiometabolic risk factors than females in NHs. ^18^

An acute health crisis can produce a harvesting effect as was suggested during the 2016 influenza A epidemics. ^16^ We did not observe a harvesting effect during the study period (ending in August 2020, i.e., three months after the end of the first wave). A similar observation was made in France after the August 2003 heatwave; 15,000 excess deaths were not followed by a lack of deaths until the following year.^28^ This finding may indicate that the deaths observed during first wave of the COVID-19 pandemic had not affected only those whose health was already so compromised that they “would have died in the short term anyway” but also compromised the health of the survivors. Another explanation might be the persistence of COVID-19 until August -the very beginning of the second wave. Lastly, COVID-19 survivors from the first wave may have been weakened and may have died afterwards.

In line with previous studies, we observed marked spatial heterogeneity in excess deaths among NH residents. ^29,23^ This heterogeneity might be due to differences in the regional distribution of SARS-CoV-2 infections (notably in north-east France), urban density, and other environmental factors. ^23,30,31^

Moreover, the pandemic and the corresponding public health responses had both positive and negative direct and indirect effects on the general population by increasing social isolation, reducing the frequency of road traffic accidents and occupational injuries, and restricting access to care for health conditions other than COVID-19. ^12,13,24,32-34^ Similarly, the pandemic and the corresponding public health responses might have had both direct and indirect effects on NHs. Although it is hard to imagine that COVID-19 had positive effects in NHs, this topic has not been studied and the part of direct and indirect effects of pandemic on excess mortality remains unknown. Our results suggest that COVID-19 had a large, direct, negative impact on excess mortality in NHs. The number of excess death estimated here (15,000) was slightly higher than the nationally reported number of COVID-19-related deaths among NH residents (14000). ^1^ The changes over time in excess deaths and the concentration of these deaths in the territorial units most strongly affected by SARS-CoV-2 also argue in favor of a direct effect. However, the presence of excess deaths in some territories barely or not impacted by the first wave of COVID-19 infection argues in favor of the presence of both direct and indirect effects. Moreover, a proportion of the deaths attributed to COVID-19 by the French national surveillance system might have another cause because both suspected and confirmed cases were recorded at the start of the pandemic.^1^ The present study had several strengths. Firstly, the nationwide setting enabled us to extrapolate our results to the entire population of NH residents. Secondly, access to reference data from the previous 6 years in the same population enabled us to smooth out year-on-year variations in mortality. However, given that the impact of influenza epidemics on NH mortality varies widely from one year to the next, it would be interesting to compare 2020’s excess deaths with each single reference year as a function of the influenza epidemic at that time.

## Conclusion

We estimated that during the first wave of the COVID-19 pandemic, 15,114 excess deaths occurred among NH residents in France. These deaths accounted for 51% of the total excess deaths in the general population. The mortality increased overall by 43%. The excess mortality was higher among males than among females (51% and 38%, respectively) and decreased with age. Excess deaths varied widely across from one territorial unit to another. We did not observe a harvesting effect during the study (up until August 30^th^, 2020).

Real-time mortality surveillance in both NHs and hospitals is critically important for monitoring epidemics and other health crises and for guiding public health decisions.^35^ The analysis of causes of death (when available) should help to differentiate between deaths directly vs. indirectly caused by COVID-19. A refined analysis is required to identify individual and environmental risk factors associated with SARS-CoV-2 infections and COVID-19 lethality. In particular, the roles of comorbidities, old age, dependency, the resident:staff ratio, the use of personal protective equipment, NH crowding, population density, and other socio-economic factors must be characterized before models of care and nursing for dependent older adults are designed.^36^

## Data Availability

The data are not available.

## Author Contributions

### Concept and design

Florence Canouï-Poitrine, Marie Laurent, Antoine Rachas

Acquisition, analysis, or interpretation of data: Florence Canouï-Poitrine, Martine Thomas, Laure Carcaillon-Bentata, Romeo Fontaine, Gaetan Gavazzi, Jean-Marie Robine, Marie Laurent, Antoine Rachas.

### Drafting of the manuscript

Florence Canouï-Poitrine, Laure Carcaillon-Bentata, Romeo Fontaine, Jean-Marie Robine, Marie Laurent, Antoine Rachas.

### Critical revision of the manuscript for important intellectual content

Florence Canouï-Poitrine, Martine Thomas, Laure Carcaillon-Bentata, Romeo Fontaine, Gaetan Gavazzi, Jean-Marie Robine, Marie Laurent, Antoine Rachas.

### Statistical analysis: Martine Thomas

Antoine Rachas and Martine Thomas had full access to all of the study data and take responsibility for the integrity of the data and the accuracy of the analysis.

## Additional contributions

the authors thank Sylvie Bastuji-Garin, MD, PhD (Univ Paris Est Creteil/APHP), Costas Danis (Santé Publique France), Sylvain Diamantis, MD, PhD (Melun Hospital), Laure Fonteneau, MD, PhD (Santé Publique France), Emmanuel Forestier (Chambery Hospital), Anne Gallay, MD, PhD (Santé Publique France), Claire Roubaud, MD, PhD (Bordeaux University and Hospital), Ayden Tahjamady MD, PhD (Cnam) Pierre Wolkenstein, MD, PhD (Univ Paris Est Creteil/APHP). These people were not remunerated for their assistance with the present study. The multidisciplinary COVID Mortality in Nursing Homes (COMONH) consortium comprises epidemiologists (Florence Canouï-Poitrine, Laure Carcaillon-Bentata, Costas Danis, Laure Fonteneau, Antoine Rachas), a statistician (Martine Thomas), geriatricians (Emmanuel Forestier, Gaëtan Gavazzi, Marie Laurent, Claire Roubaud), a demographer (Jean-Marie Robine) and an economist (Roméo Fontaine). We thank David Fraser, PhD for manuscript editing.

